# Exploring the use of UmbiFlow^™^ to assess the impact of heat stress on fetoplacental blood flow in field studies

**DOI:** 10.1101/2022.03.31.22273092

**Authors:** Ana Bonell, Valerie Vannevel, Bakary Sonko, Nuredin Mohammed, Ana M. Vicedo-Cabrera, Andy Haines, Neil S Maxwell, Jane Hirst, Andrew M Prentice

**Affiliations:** Medical Research Council Unit The Gambia at London School of Hygiene and Tropical Medicine, Banjul, The Gambia; Centre on Climate Change and Planetary Health, London School of Hygiene and Tropical Medicine, London, UK; Maternal and Infant Healthcare Strategies Unit, SAMRC, Pretoria, South Africa; Department of Obstetrics and Gynaecology, University of Pretoria, Pretoria, South Africa; Research Centre for Maternal, Fetal, Newborn & Child Healthcare strategies, University of Pretoria, Pretoria, South Africa; Institute of Social and Preventive Medicine, University of Bern, Bern, Switzerland; Oeschger Center for Climate Change Research, University of Bern, Bern, Switzerland; Department of Public Health, Environment and Society, Department of Population Health, London School of Hygiene and Tropical Medicine, London, UK; Environmental Extremes Laboratory, University of Brighton, Eastbourne, UK; Nuffield Department of Women’s and Reproductive Health and the George Institute for Global Health, University of Oxford, Oxford, UK

**Author notes:** Corresponding author: Ana Bonell, Medical Research Council Unit The Gambia at London School of Hygiene and Tropical Medicine, MRC Fajara, Atlantic Boulevard, Banjul, The Gambia. joint last author (Jane Hirst, Andrew M Prentice).

**Keywords:** heat, pregnancy, fetoplacental circulation, climate change, Africa

## Abstract

**Objective:** To evaluate the impact of heat stress on umbilical artery resistance index (RI) measured by UmbiFlow™ in field settings and the implications for pregnancy outcomes.

**Methods:** This feasibility study was conducted in West Kiang, The Gambia, West Africa; a rural area with increasing exposure to extreme heat. We recruited women with singleton fetuses who performed manual tasks (such as farming) during pregnancy. The umbilical artery RI was measured at rest, during and at the end of a typical working shift in women ≥ 28 weeks’ gestation. Adverse pregnancy outcomes (APO) were classified as stillbirth, preterm birth, low birth weight, or small for gestational age, and all other outcomes as normal.

**Results:** A total of 40 participants were included; 23 normal births and 17 APO. Umbilical artery RI demonstrated a nonlinear relationship to heat stress, with indication of a potential threshold value for placental insufficiency around 32ºC by universal thermal climate index. Preliminary evidence suggests the fetoplacental circulation response to heat stress differs in APO versus normal outcome.

**Conclusions:** The Umbiflow™ device proved to be an effective field method for assessing placental function. Dynamic changes in RI may begin to explain the association between extreme heat and APO.

**Funding:** The Wellcome Trust (216336/Z/19/Z)

**Synopsis:** Extreme heat exposure is increasing and a low-cost umbilical artery doppler device, UmbiFlow™, can aid understanding of fetoplacental function under heat stress conditions.

## Introduction

With the ongoing climate crisis, global extreme heat exposure is progressively increasing with, for example, 30% of the world’s population already exposed for 20 or more days annually to levels of heat sufficient to cause excess mortality and up to 74% are predicted to be exposed by 2100.(1) Sub-Saharan Africa (SSA), South and South East Asia have been identified as regions at high risk of climate change related extreme weather events, despite contributing almost nothing to the problem.(2) In The Gambia, West Africa, extreme heat, defined as above the 90% centile compared to the average temperature for that region (>39.4°C), occurred on average for 50 days per year, from 2016-2019 (from local weather station data). The double burden of deadly heat exacerbated by climate change and existing health inequalities make this a critical location to study.

The burden of adverse pregnancy outcomes (APOs) are mainly felt in low middle income countries (LMICs), for example an estimated 15 million preterm births (PTB) occur per year, with greater than 80% occurring in Asia and SSA.(3) PTB is linked to high rates of both perinatal mortality (the cause of up to 24% of SSA neonatal deaths) and morbidity with long-term implications.(4) Triggers for preterm labour are complex and multifactorial, but recent environmental epidemiological studies demonstrate that maternal exposure to extreme heat increases the risk of PTB.(5, 6) Stillbirths, a neglected tragedy are again mainly felt in LMICs, with increasing rates and have also been linked to extreme heat exposure.

The impact of heat on pregnancy depends on the intensity, duration and exposure window. First trimester exposure leads to increased embryonic death, cardiac and neurological anomalies.(7) In the second and third trimester, maternal exposure to ambient heat has been shown to increase the risk of PTB, stillbirths and low birth weight (LBW) in multiple settings.(6, 8, 9) Despite strong environmental epidemiological evidence of this linkage, there remains limited understanding of the pathophysiological mechanism associated with these poor outcomes.(10) One of the proposed hypotheses is that thermoregulatory changes to blood flow prioritise heat loss through cutaneous vasodilation over other homeostatic mechanisms. For example, in non-pregnant individuals during exertional heat strain, mesenteric and renal blood flow can be reduced to such an extent that gut permeability or acute kidney injury may occur.(11) In pregnancy, where blood flow to the uterus and placenta depends on cardiac output, with no autoregulation, there is evidence from animal studies that this occurs,(12) but human studies are lacking.(10) However, placental insufficiency is implicated in the pathophysiological mechanisms of stillbirth, preterm birth and intrauterine growth restriction.(13) Heat stress could potentially impact on fetal wellbeing if the placenta is unable to buffer the effects of the reduction in blood flow leading to transient placental insufficiency. Thus, identification of individuals in whom blood flow to the uterus and placenta is reduced during a heat stress event, could allow preventive measures to be taken.

Direct measurement of blood flow to the placenta through the uterine arteries can be challenging as it requires highly specialised non-portable equipment in conjunction with fluid dynamic modelling.(14) However, the umbilical artery doppler waveform gives an indication of the fetoplacental circulation function, and so indicates how effectively the fetus is receiving oxygen, nutrients and removing waste products, and can be used as a surrogate for direct blood flow measurement. The UmbiFlow™ device, a low-cost portable continuous-wave doppler device was designed and developed in South Africa and has been validated for use to identify placental insufficiency based on the resistance index (RI) of the umbilical artery, with accuracy comparable to commercial units.(15, 16) It has not yet been used to explore dynamic changes in the RI under different physiological conditions. We hypothesise that underlying placental problems that may then lead to APO will alter the effect of heat on umbilical artery RI, with those who have APO being more likely to have placental insufficiency under heat stress. Therefore, the following study objectives were defined:

- determine if UmbiFlow™ identifies a change in umbilical artery resistance index under heat stress; and
- determine the sample size that would be needed to definitively test the association between changes in umbilical artery RI under heat stress and adverse pregnancy outcomes.
- determine the practical considerations needed to use UmbiFlow™ in the field.

## Materials and Methods

This feasibility study was part of a larger prospective cohort study on heat strain in pregnant subsistence farmers and the physiological impact on their fetuses.(10) The study was approved by the Gambia government/MRC Joint ethics committee and the London School of Hygiene and Tropical Medicine Ethics Advisory Board (ref: 16405) in accordance with the Declaration of Helsinki (2013).

Briefly, pregnant women living in West Kiang, The Gambia, participated in an observational cohort study of maternal heat strain and the assessment of the dynamic changes in maternal and fetoplacental blood flow during a day of field work.(17) Participants were eligible if they were singleton pregnancies, undertook farming tasks during pregnancy and did not suffer with pre-eclampsia or eclampsia at the time of recruitment. Gestational age was determined by last known menstrual period when known, or biparietal diameter on ultrasound scan before 28 weeks’ gestation by a trained sonographer when unknown. The feasibility study visits occurred in those with gestational age ≥ 28 weeks, and during their usual farming activity. External environmental conditions (air temperature, relative humidity, solar radiation, wind speed) were measured hourly using the HT200: Heat Stress WBGT Meter, Extech® and the Extech® AN100 thermo-anemometer, NH, USA. Two thermal indices were calculated from these measures – the Wet Bulb Globe Temperature (WBGT) and the Universal Thermal Climate Index (UTCI). These are composite measures of thermal stress taking into account heat, humidity, solar radiation and wind speed.(18)

UmbiFlow™ measures the blood flow velocity in the umbilical cord and calculates the RI = (systolic velocity – diastolic velocity)/systolic velocity. The hand-held probe attaches to a laptop/tablet, signal processing occurs within the specialised software to give both a waveform and audible umbilical artery blood flow. Validated reference values by gestational age indicate if the RI is within normal, intermediate or high-risk range. On a single occasion for each subject the RI was measured at baseline in an airconditioned environment with the participant supine at rest, and with abdominal lateral tilt, and then at two time points during her working day. At each time point, two measurements were taken, assessed for quality (signal quality assessed by expert trained by the South African team), mean values taken when good/moderate quality and discarded if poor quality. The risk category (low risk, intermediate risk, high risk) based on normalised curves for the Umbiflow™ were recorded at each reading as well as the exact value of the RI. APOs were defined as follows: stillbirth = pregnancy > 20 weeks’ gestation where the baby was born dead; PTB = live birth prior to completion of 37 weeks’ gestation; LBW = birth weight ≤ 2.5 kg; small for gestational age (SGA) = birth weight < 10% expected at gestational age based on Intergrowth-21 standardised curves.

All analyses were performed in R version 4.1.0. Descriptive characteristics are presented as mean +/- SD or median (IQR) by outcome, depending on distribution. The relationship between UTCI, fetal heart rate and umbilical artery RI were explored using linear and non-linear models. Non-linear models were tested across different spline definitions and different knots placed at the median and 90^th^ percentile. The lowest AIC was used to determine best model fit. Change in fetal heart rate (FHR) by UTCI was best explained by a linear model. RI z-score or change in RI by UTCI was best explained by a non-linear model with a cubic spline with one knot at the median.

A multilevel linear regression model, with individual as random effect, of the association between umbilical artery RI z-score and heat stress was explored both with and without cubic splines and then stratified by APO with the best fit determined by AIC. The final model is shown:

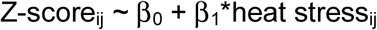

z-score = umbilical artery RI z-score for individual *i* at time *j* heat stress = UTCI for individual *i* at time *j*

Multilevel model assumptions were assessed by examining normality of residuals and performing Levene’s Tests for homogeneity of variance. The simr package was used to run a simulation-based power analysis on the multilevel model to give estimations of sample size requirements to detect a difference in umbilical artery RI z-score under heat stress in those with APO.

## Results

Full umbilical artery doppler was completed on 40 participants the field. Out of these 40 participants, 17 had APO and 23 did not. Of those with APO, 3 suffered with stillbirths, 7 delivered preterm (spontaneously), 6 were LBW and 8 had SGA. Descriptive characteristics of all participants are presented in Table 1, with detailed description of those with stillbirths in Table 2.

**Table 1:**
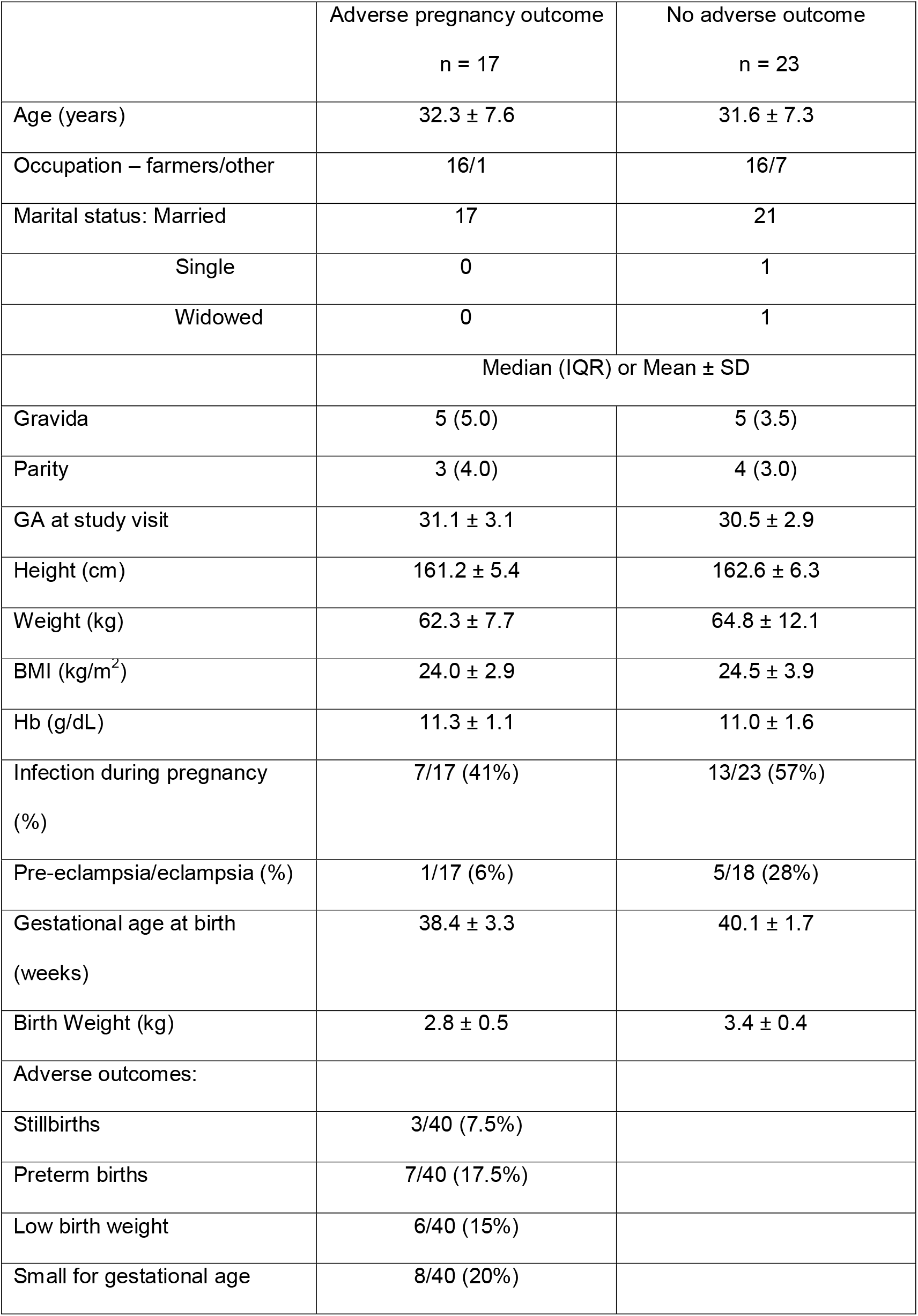
Demographic, social, obstetric and anthropometric characteristics of those with adverse pregnancy outcomes and those without.

**Table 2:** details of participants who had stillbirths.

| Case A | Case B | Case C |
| --- | --- | --- |
| Maternal age = 36-40 yrs | Maternal age = 41-45 yrs | Maternal age = 26-30 yrs |
| Previous stillbirths | No previous stillbirth | No previous stillbirth |
| GA at visit = 31+6 | GA at visit = 34+5 | GA at visit = 32+1 |
| **High RI at baseline** | **Low RI at baseline** | **Low RI at baseline** |
| **High RI during work** | **High RI during work** | **Low RI at baseline** |
| **High RI after work** | **High RI after work** | **Low RI at baseline** |
| Referred for urgent care | Referred for urgent care | Normal care |
| GA at delivery = 37+1 | GA at delivery = 40+1 | GA at delivery = 42+2 |
| Stillbirth | Stillbirth | Likely intrapartum death |

Environmental conditions and physiological parameters at baseline, during the work shift and at the end of the work shift are presented in Table 3. All participants were exposed to “extreme heat stress” (based on the UTCI value), which has been shown to increase risk of mortality in other populations and settings.(19) Average physical energy expenditure for the working shift was equivalent to moderate intensity exercise such as a brisk walk.(20) There was no significant difference between working environmental conditions or estimated energy expenditure in those who went on to have an APO compared to those who did not.

**Table 3:** Mean +/- SD of environmental conditions, maternal tympanic temperature, fetal heart rate and umbilical artery resistance index.

|  | Baseline |  | Mid-way |  | End of shift |  |
| --- | --- | --- | --- | --- | --- | --- |
|  | APO | No APO | APO | No APO | APO | No APO |
| UTCI (°C) | 23.1 ± 1.3 | 22.1 ± 2.2 | 34.2 ± 4.0 | 33.6 ± 4.2 | 34.1 ± 3.2 | 34.5 ± 2.7 |
| WBGT (°C) | 19.3 ± 1.0 | 18.6 ± 1.8 | 27.3 ± 4.1 | 27.2 ± 4.5 | 27.3 ± 3.6 | 27.7 ± 2.7 |
| Air temp (°C) | 24.0 ± 1.5 | 22.9 ± 2.0 | 34.2 ± 3.6 | 33.9 ± 4.1 | 34.4 ± 3.2 | 34.8 ± 3.0 |
| T<sub>tym</sub> (°C) | 36.9 ± 0.2 | 36.9 ± 0.2 | 37.2 ± 0.4 | 37.1 ± 0.3 | 37.1 ± 0.3 | 37.3 ± 0.3 |
| PAEE<br>(kcal/kg/hr) | - | - | - | - | 3.1 ± 0.9 | 3.1 ± 0.7 |
| FHR (b.min<sup>-1</sup>) | 128.4 ± 8.4 | 126.4 ± 7.8 | 142.2 ± 14.4 | 147.3 ± 7.1 | 143.7 ± 11.8 | 144.9 ± 11.5 |
| RI | 0.68 ± 0.10 | 0.65 ± 0.08 | 0.69 ± 0.10 | 0.67 ± 0.05 | 0.66 ± 0.05 | 0.65 ± 0.07 |
| z-score | 0.74 ± 1.59 | 0.43 ± 1.23 | 0.964 ± 1.58 | 0.52 ± 0.74 | 0.51 ± 0.89 | 0.23 ± 1.13 |

UTCI = universal climate thermal index; WBGT = wet bulb globe temperature; T<sub>tym</sub> = tympanic temperature; PAEE = physical activity energy expenditure; FHR = fetal heart rate; RI = umbilical artery resistance index.

Fetal heart rate demonstrated a linear relationship with heat stress, giving an increase of 12 beats per minute by each 10°C UTCI increase (Figure 1A). However, there was no clear linear or nonlinear relationship between fetal heart rate and maternal tympanic temperature. Change in RI from cool baseline to working conditions reduced with increasing heat stress exposure up to 32°C UTCI and then appears to begin to increase with rising heat stress (Figure 1B). A similar finding was seen with WBGT. The change in RI stratified by APO is given in Figure 2A&B and shows that in those with APO there was an increase in RI with heat stress exposure.

**Figure 1:**
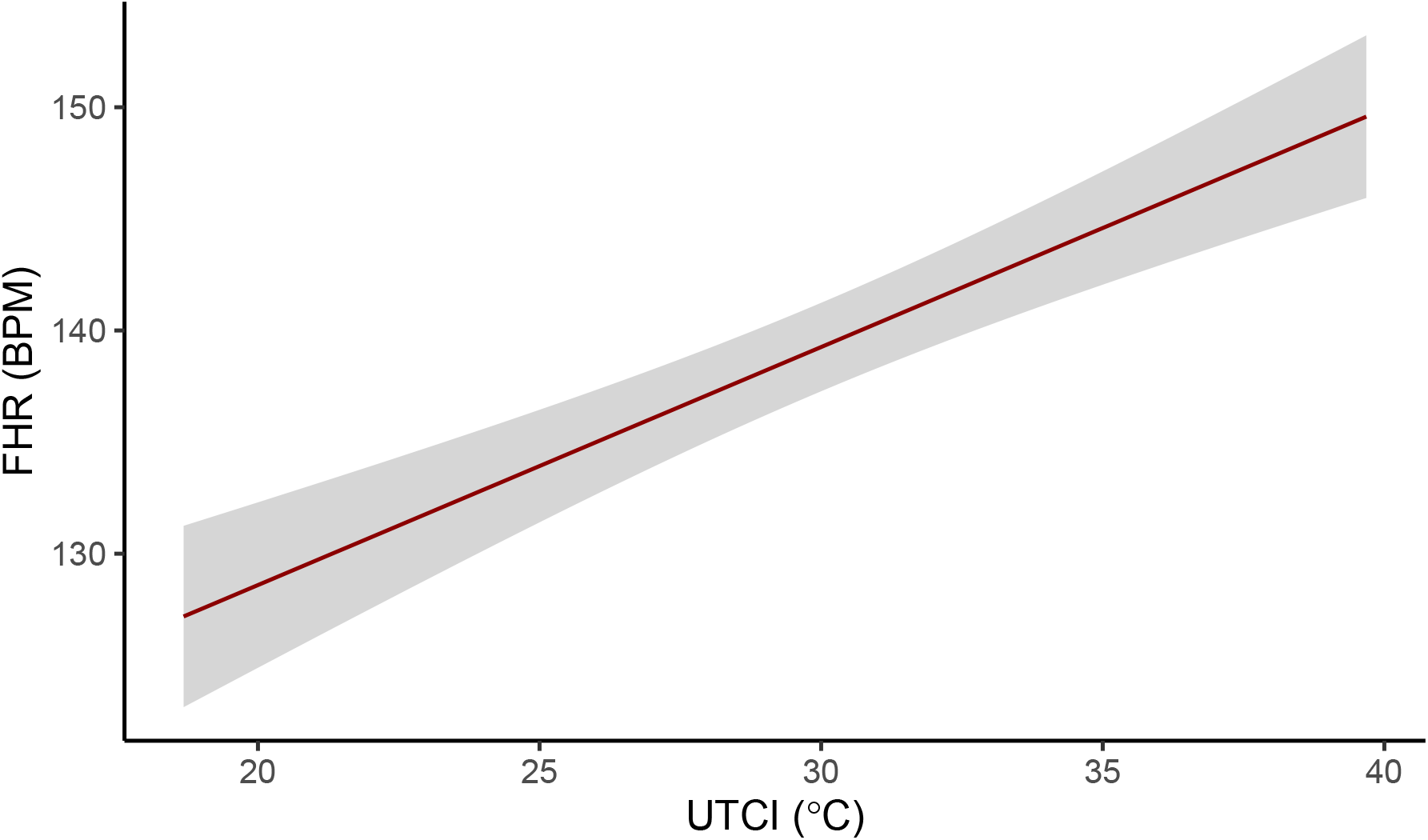
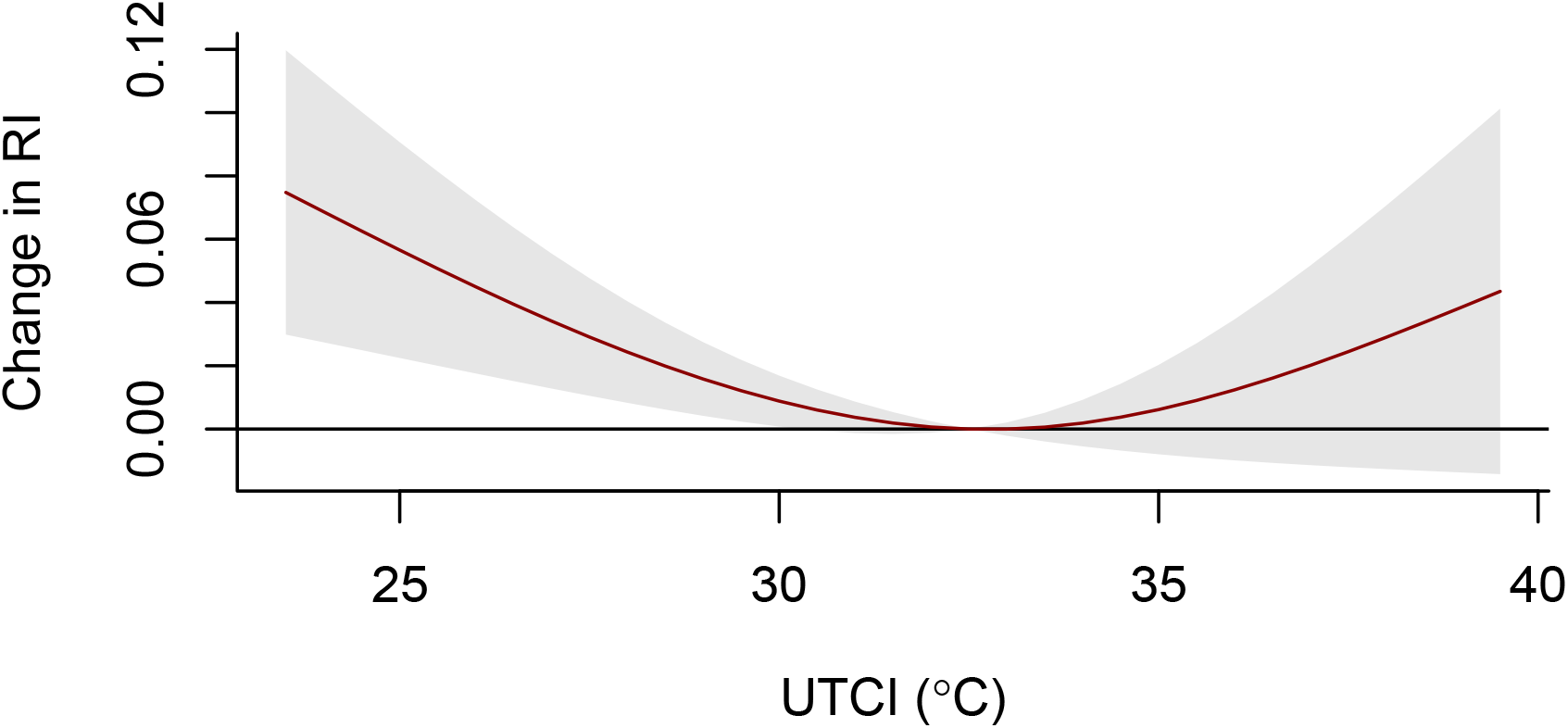
Association between FHR (A), change in umbilical artery RI (B) and heat stress (UTCI).

**Figure 2:**
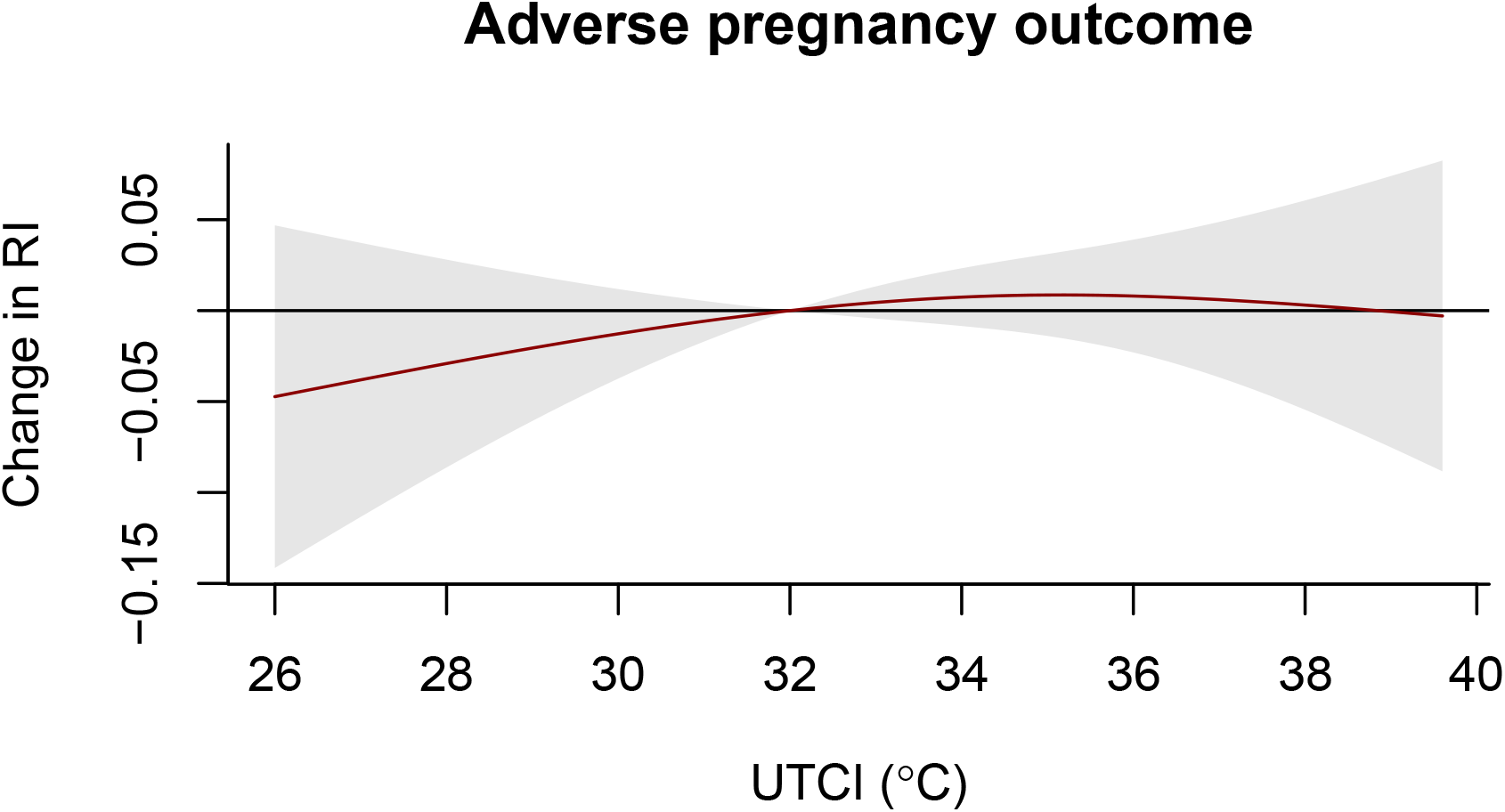
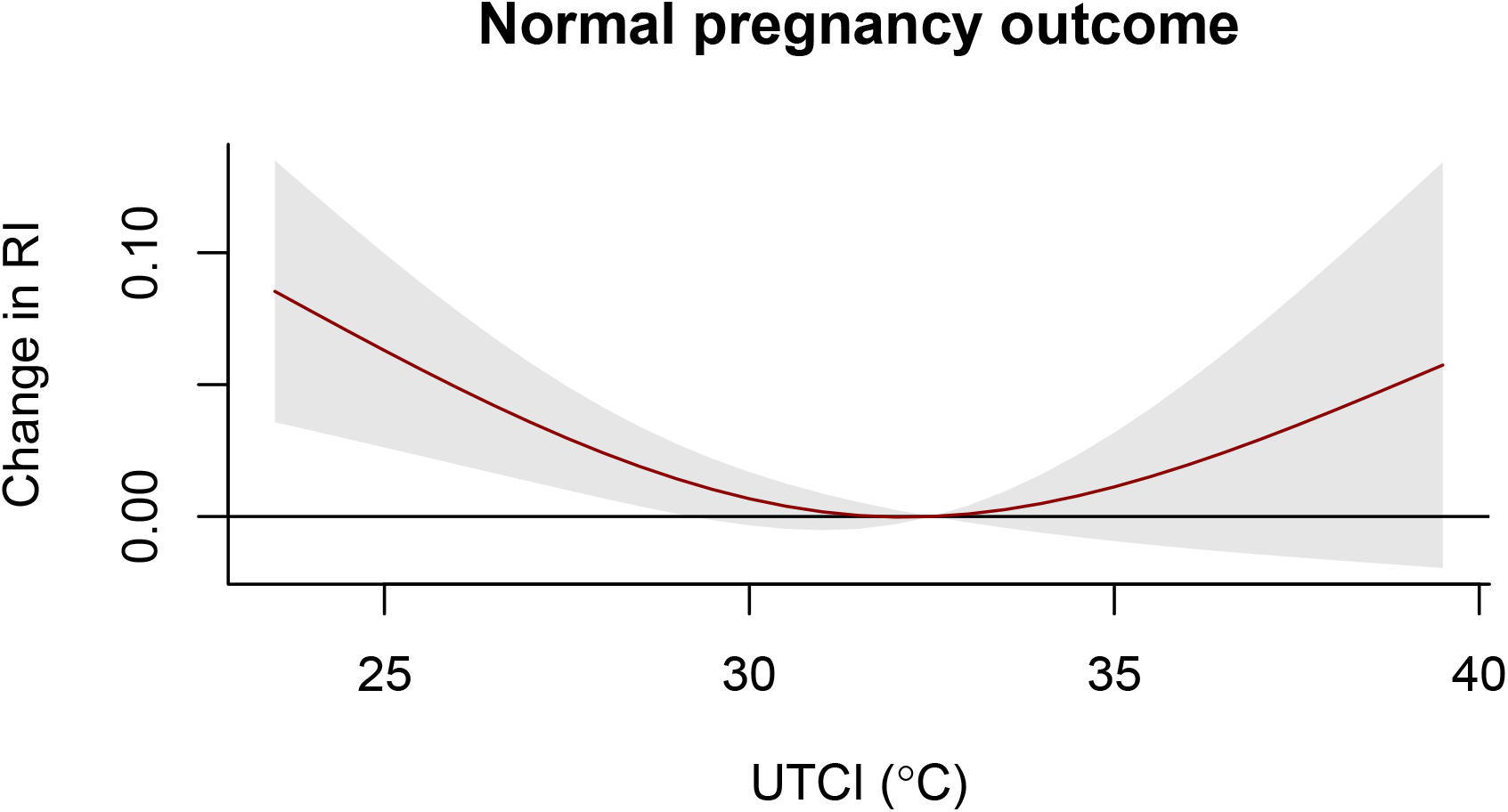
Association between change in umbilical artery RI under heat stress (UTCI) in those with APO (A) and normal birth outcomes (B).

Our model estimates showed an increase of 0.02 (95% CI: -0.25;1.11) in umbilical artery RI z-score in those with APO with each 1 degree increase in UTCI. Model diagnostics for normality of residuals and homogeneity of variance (using Levene’s Test) did not indicate gross violation of model assumptions. Using the output from the multilevel model, a simulation-based power calculation using simr package was run to identify the sample size required to determine the relationship between umbilical artery z-score and heat stress in those with APO with a power of 80% and an alpha of 0.05. The full output is shown in Table 4 and Figure 3, giving a sample size estimate of 997 individuals to reach statistical significance.

**Table 4:** Power calculation predictor for determining association between umbilical artery resistance index z-scores under heat stress in those with adverse pregnancy outcomes.

| Highest UTCI exposure (°C) | Power to detect effect size (%) | 95% CI | Sample size |
| --- | --- | --- | --- |
| 21.70 | 0 | 0.0;0.4 | 93 |
| 23.14 | 0 | 0.0;0.4 | 249 |
| 23.74 | 0 | 0.0;0.4 | 375 |
| 25.83 | 8.1 | 6.5;10.0 | 531 |
| 32.39 | 41.7 | 38.6;44.8 | 686 |
| 33.27 | 68.5 | 65.5;71.4 | 841 |
| 34.92 | 91.1 | 89.2;92.8 | 997 |
| 36.82 | 99.2 | 98.4;99.7 | 1186 |
| 37.46 | 99.9 | 99.4;100.0 | 1311 |
| 39.68 | 100 | 99.6;100.0 | 1468 |

**Figure 3:**
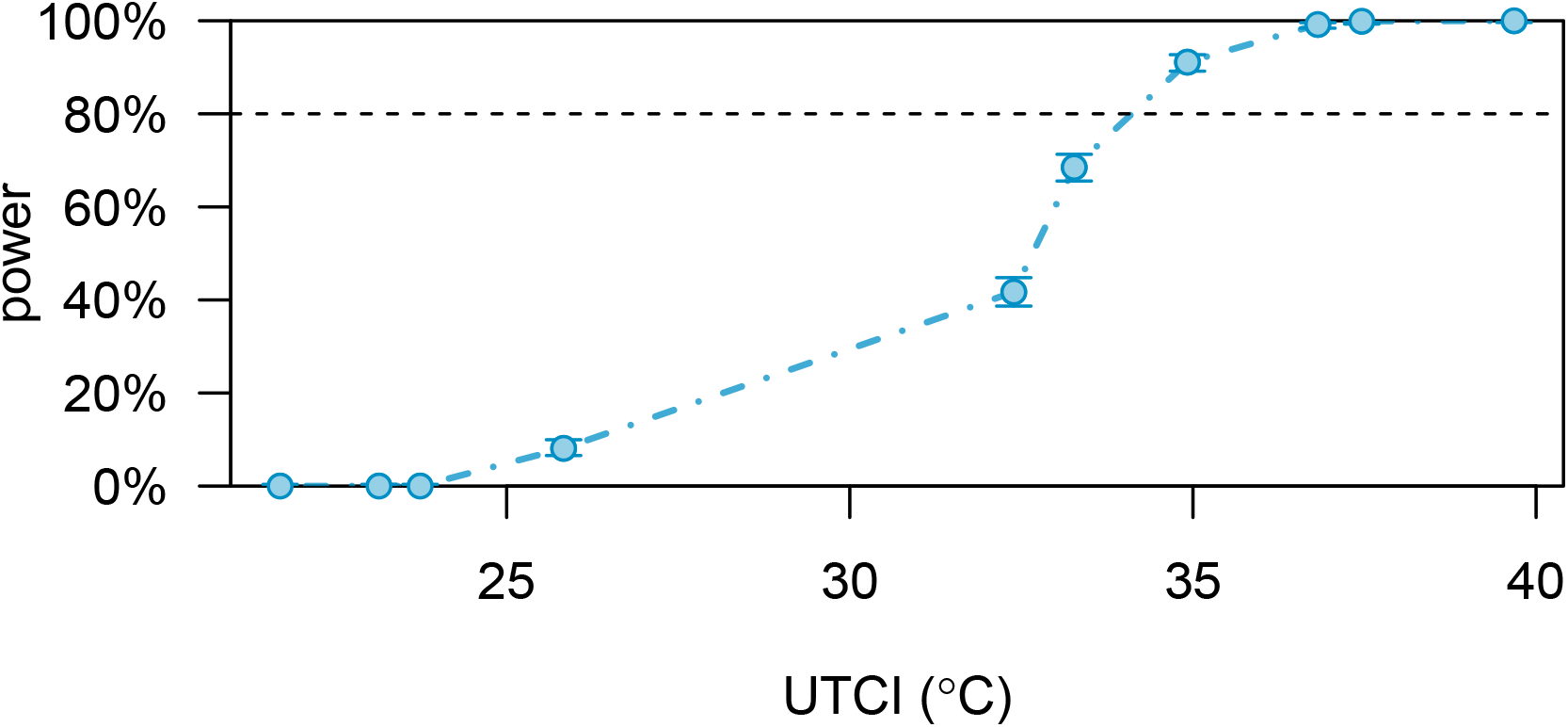
Simulation-based power calculation to demonstrate a significant association between umbilical artery z-scores and heat stress (UTCI) in those with APO.

## Discussion

We show that the measurement of umbilical artery doppler in field conditions is possible and shows promising evidence of potentially enhancing the understanding of the fetoplacental circulation response to heat stress. Under heat stress conditions below 32°C UTCI there was a reduction in the umbilical artery RI from baseline, which would indicate increased blood velocity within the fetoplacental circulation. However, above this temperature threshold there appears to be a trend towards increasing RI which would indicate insufficiency in the fetoplacental circulation. The response to heat stress appears to be different in those individuals that went on to have an APO, however we were not powered to determine this with statistical significance. Based on these preliminary findings, a simulated sample size of 997 pregnant women with APO, would reach statistical significance to demonstrate the relationship between umbilical artery RI z-score and heat stress.

The UmbiFlow™ device is highly suited to field work, being light, compact and compatible with any PC laptop with Windows 7 or 10 installed and requiring minimal training. Practical considerations for use in the field included ensuring comfortable and private area to scan - which was provided by local vegetation or screens; protection from extreme weather – provided by portable shade/rain protector; and need for accurate gestational age to calculate RI z-scores.

There are few studies exploring the impact of heat on uterine or placental blood flow. A study from Sweden on sauna use (20 mins at 70°C) in late pregnancy found a reactive increase in fetal heart rate, but no change in umbilical artery blood flow.(21) This study is not immediately translatable to other settings due to both the inactivity and the extreme heat, but could be reassuring in terms of short bursts of unavoidable extreme heat exposure. Other studies have mainly focused on thermoregulation in pregnancy and there are several studies with encouraging evidence that thermoregulation is not compromised.(22, 23) Although there is clear evidence that moderate intensity exercise is of benefit in pregnancy,(24) these studies are in temperate conditions and so not transferable to our setting. Additionally in extreme cases (Olympic athletes exercising at > 90% maximum maternal heart rate) there can be compromised fetal wellbeing.(25) This extreme physiological strain may be similar to that experienced under extreme heat and warrants further investigation.

The study has several limitations. The sample size was reduced due to the covid-19 pandemic halting all field work activity from March 2020, limiting the scope of analysis available. Maternal core temperature could not be measured in the field (impractical to use rectal thermometer and lack of evidence on safety for core telemetry pills) and therefore the less accurate and less precise tympanic temperature was measured. Additionally, pregnancy and neonatal outcomes in the general population of The Gambia are worse than the global average which may impact on generalisability of the findings globally but could be reasonably representative of a rural SSA population. This study comes at a time where extreme heat exposure is becoming a reality for much of the global population. Despite this, those most commonly experiencing these extreme conditions are often missing from the medical literature. This study is set in a rural African setting, with a population that can be difficult to access but are often exposed to extreme environmental conditions. By exploring ways to improve the understanding of pathophysiological mechanisms in a real-life setting we highlight the need for future work. The simulation-based sample size calculations give an estimate of the sample size and conditions needed to progress understanding of this using the Umbiflow™ device. However, without expanding the work to include several key areas the impacts of this research will have little meaning to this population. Identifying at risk women will not be beneficial without clear management options to reduce the risk of these adverse outcomes. Health system strengthening in both facilities and human capacity in dealing with maternal health are urgently needed especially in the face of the growing climate crisis and resultant impacts on healthcare. Additionally, heat exposure is increasing and evidence-based, effective, realistic, pragmatic and sustainable interventions for cooling both individuals and their environment are urgently needed.

## Data Availability

Anonymised data will be made available on reasonable request from the corresponding author.

## Author contributions

AB: conceptualization, methodology, formal analysis, writing original draft.

VV: methodology, validation, editing.

BS: software, data curation, editing.

NM: formal analysis, editing.

AVC, AH, NM, JH, AP: conceptualization, methodology, supervision, editing

## Data availability

Anonymised data will be made available on reasonable request from the corresponding author.

